# Serial visualization of Glucose-insulin metabolism in thin young Indians with prediabetes reveals early failure of beta cell secretion in relation to decreasing insulin sensitivity

**DOI:** 10.1101/2022.10.15.22281121

**Authors:** Pradeep Tiwari, Sanat Phatak, Souvik Bandopadhyay, Tavpritesh Sethi, Caroline HD Fall, Chittaranjan Yajnik

## Abstract

**Background & Objective:** Defective beta cell function in relation to impaired insulin sensitivity results in glucose intolerance. There are few studies documenting the lifecourse evolution of this relationship. The Pune Maternal Nutrition Study (PMNS) longitudinal birth cohort offered the opportunity to document these parameters from childhood in young, rural prediabetic participants and compare them to normal glucose tolerant (NGT).

**Methods:** PMNS subjects were classified as NGT or Glucose intolerant according to their OGTT results at 18 years of age. Insulin Sensitivity (HOMA-S) and β-cell function (HOMA-β) were estimated at 6,12 and 18 years. Their inter-relationship was estimated using HOMA-β as a nonlinear function of HOMA-S, separately for NGT and Glucose intolerant individuals at 6,12 and 18 years. Rates of change of HOMA-S and HOMA-β were estimated using a linear mixed effect model and visualized using LOESS plots.

**Result:** Of 619 participants, 177 had glucose intolerance at 18 years of age. A nonlinear hyperbolic relationship between HOMA-S and HOMA-B was observed at all time points. There was a progressive fall in HOMA-S and rise in HOMA-B with increasing age. Glucose intolerant participants had lower HOMA-B for all levels of HOMA-S as compared to NGT, manifesting as shift towards the origin in the hyperbolic curve.

**Conclusion:** We provide evidence for early life dysregulation in glucose insulin metabolism leading to pre-diabetes at 18 years of age. Prediabetic individuals started with lower beta cell function and lower insulin sensitivity from an early age. Diabetes prevention should start from early life.

## Introduction

The action of insulin in efficient glucose metabolism is primarily governed by two parameters: insulin sensitivity (the ability of peripheral tissue to respond to insulin) and the amount secreted. [1] Bergman first described the relationship under normal physiological conditions, where a decrease in insulin sensitivity is compensated by an increase in insulin secretion to maintain glucose disposal.[2] Their non-linear relationship plots as a rectangular hyperbolic curve such that at any point of time, their product (disposition index) remains constant.

Type 2 diabetes ensues when there is a reduction in both sensitivity and secretion;[1] a reduction in one parameter alone is not enough to cause hyperglycemia. There has been ongoing debate about the ‘first’ hit in the pathogenesis, with existing arguments for both an early reduction in sensitivity [3,4] and secretion [5,6]. The relative contribution of both these probably varies in different individuals and situations. [1]

India is facing a burgeoning diabetes epidemic [7]; Indians get diabetes at a younger age and lower BMI compared to the western population [8]. It was traditionally thought that Indians have lower insulin sensitivity [9], but recent evidence suggests early-life insufficiencies in insulin secretion. [10] Majorities of these reports are based on cross-sectional studies or short prospective studies in adults. We had the opportunity to examine the evolution of insulin function in the Pune Maternal Nutrition Study (PMNS), a rural Indian birth cohort setup in 1993 with serial metabolic measurements through childhood. One-third of the participants had glucose intolerance at 18 years of age. [10]. We tested the nature of the relationship using HOMA models, and plotted trajectories to depict the evolution of glucose-insulin metabolism in this cohort in a visually intuitive manner.

## Methods

### Cohort

The PMNS was established in 1993 in six villages near Pune, Western India, to prospectively study associations of maternal nutritional status with fetal growth and later diabetes risk in the offspring. [11] Briefly, Married women were followed up and those who became pregnant (F0 generation) were recruited into the study. Their children (F1 generation, participants in this study) were followed up at birth, 6, 12 and 18 years of age. Participants arrived at the Diabetes Unit, KEM Hospital, the evening before the day of blood sampling, had a standard dinner, and fasted overnight. In the morning, a fasting blood sample was collected. At 6 years, an oral glucose tolerance test (OGTT) was performed, using 1.75g/kg of anhydrous glucose, followed by further samples at 30 and 120 minutes. At 12 years, only a fasting sample was collected. At 18 years a full OGTT (75g anhydrous glucose) was repeated. Glucose was measured by the glucose oxidase/peroxidase method, and specific insulin by ELISA. Insulin sensitivity (HOMA-S) and beta cell function (HOMA-β) were calculated using data from the fasting samples and the iHOMA2 website. [https://www.phc.ox.ac.uk/research/technology-outputs/ihoma2, last accessed August 2019]. At each timepoint, the disposition index was calculated as a product of insulin sensitivity (HOMA-S) and β-cell function (HOMA-β).

The study was approved by village leaders and the KEM Hospital Research Centre Ethics Committee. Children under 18 years of age gave written assent and their parents gave consent; they gave written consent at 18 years.

### Definitions

Using American Diabetes Association criteria, participants at 18 years were classified as normal glucose tolerance (NGT) if their Fasting Plasma Glucose < 100 mg/dL and 2-h Plasma Glucose < 140 mg/dL), Glucose intolerant if Fasting Plasma Glucose: 100 - 125 mg/dL and/or 2-h Plasma Glucose: 140 mg/dL - 199 mg/dL, and diabetes if Fasting Plasma Glucose >126 mg/dL or 2H plasma glucose > 200. [12]

### Analysis

The relationship between insulin sensitivity and β-cell function was plotted for the cohort at 6,12 and 18 years of age. LOESS models were fitted to estimate the mean curves of HOMA-β as a function of HOMA-S in NGT and Glucose intolerant (classified at 18 years). To determine if NGT and Glucose intolerant mean curves were statistically different in their slope and intercept, data were log transformed and a model was built consisting of interaction terms of HOMA-S and glucose tolerance status (Categorical, binary values; NGT, Glucose intolerant) as predictors to estimate HOMA-β. Slope and intercepts of NGT and Glucose intolerant groups were compared in the interaction model. The values on the mean curves were considered equivalent to the ‘disposition index’ (HOMA-β adjusted for HOMA-S). We compared these values between NGT and Glucose intolerant groups from childhood to young adult age.

### Trajectories of insulin sensitivity and β-cell function in NGT and Glucose intolerant

The temporal evolution of the relationship between insulin sensitivity and β-cell function from childhood (6 years) to young adult age (18 years) was plotted separately in the NGT and Glucose intolerant groups. The points on the mean curve can also be considered to be equivalent to the disposition index (HOMA-β adjusted for HOMA-S). The trajectory of this ‘disposition index’ was visualized as the plot of median values of HOMA-S and HOMA-β for each group serially over time. Mean smoothed trajectory (along with the 95% confidence interval) of disposition index, HOMA-S and HOMA-β with age (childhood to young adulthood) was estimated using LOESS regression.

The rates of change of HOMA-β, HOMA-S and disposition index (HOMA-β*HOMA-S) with age in NGT and Glucose intolerant groups were estimated using a linear mixed effect model, accounting for variations arising due to repeated measures for each participant. All the above analyses were performed on the cohort as a whole, and then repeated separately in male and female data.

### Comparison of predictive ability of Disposition Index, HOMA-S and HOMA-β for glucose intolerance

To determine how HOMA-S, HOMA-B and disposition index performed in prediction of 18-year glucose intolerance, we built binomial logistic models to classify NGT & Glucose intolerant groups for each time point. The data were partitioned into training (80%) and test set (20%). We used HOMA-S, HOMA-B and Disposition index (DI) separately as predictors after adjusting for age and sex. ROC curve analysis was performed on the test set to compare the AUC for DI, HOMA-S and HOMA-B each at each time point.

For all analyses, we used the R statistical programming language. Plots were made in the ggplot2 library, while linear mixed effect models were implemented using the lme4 library.

## Results

### Description of cohort at 18 years

The analysis included 619 participants (336 males) with complete data. (Table 1) Median height in men and women were 169.77 cm and 157.15 cm respectively. Median BMI was 19.03 kg/m2 in men and 18.06 kg/m2 in women participants. 41% of men and 57% of women were underweight (BMI <18.50 kg/m^2^). 8 % of men and 4% of women were overweight/obese (BMI ≥25 kg/m^2^). A total of 177 (28%) were glucose intolerant at 18 years of age (38% men and 18% women). Glucose intolerant men had higher BMI compared to NGT, though still classified as normal BMI, and a third were still underweight; there was no difference in BMI in women who were NGT or glucose intolerant, and two-thirds of glucose intolerant women were underweight.

### Relationship between insulin sensitivity and β-cell function in NGT vs Glucose intolerant

We observed that a hyperbolic relationship exists between insulin sensitivity (HOMA-S) and β-cell function (HOMA-B) in both NGT and glucose intolerant individuals at 6,12 and 18 years each [Figure 1A, B, C]. Hyperbolic curve for the glucose intolerant group was found to be shifted towards the origin (left and downwards) as compared to NGT. The separation between the NGT and Glucose intolerant curve was most prominent at 18 years, though a similar trend was observed at 12 and 6 years of age. The change in slope and intercepts of NGT and Glucose intolerant curves (log converted) were not found to be significantly different at any time point.

**Figure 1A:**
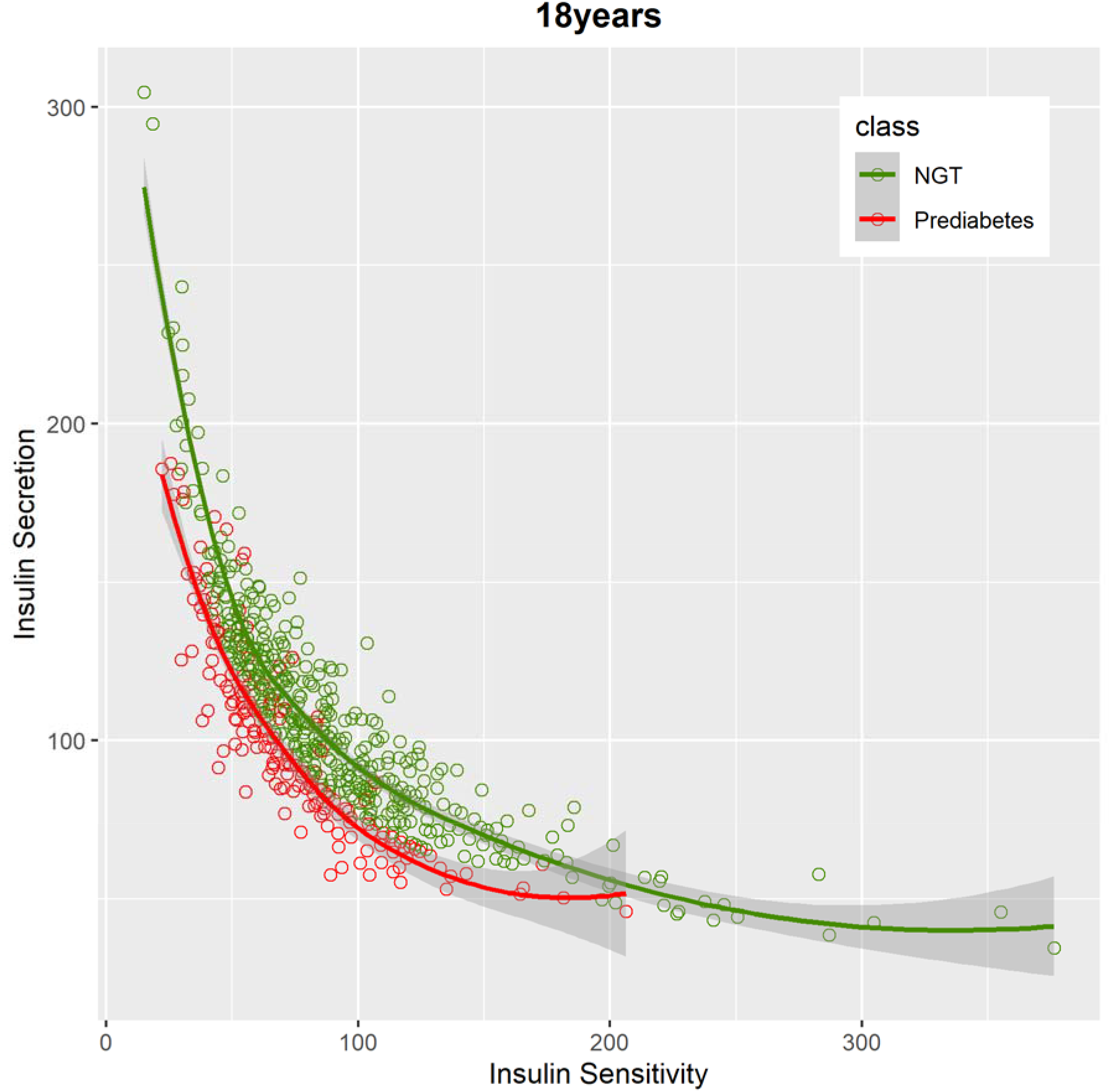
Relationship between Insulin Sensitivity (HOMA-S) and insulin secretion (HOMA-B) for both Prediabetes and Normal Glucose tolerance (NGT) individuals (18 years curve). A hyperbolic relationship was found to exist between HOMA-S and HOMA-B in both NGT and Prediabetes individuals. Curve for prediabetes was shifted more toward the left and origin.

**Figure 1B:**
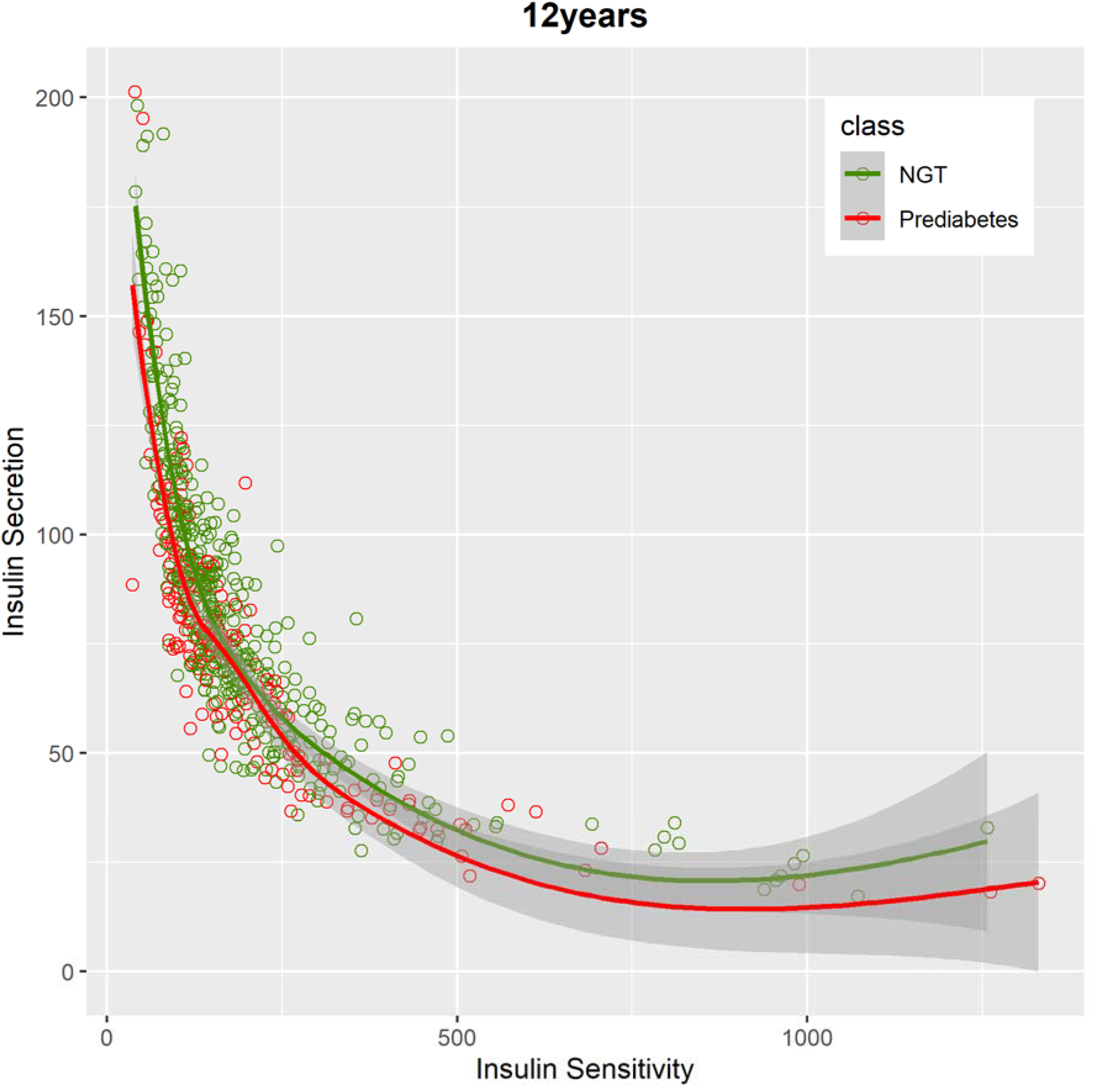
Relationship between Insulin Sensitivity (HOMA-S) and insulin secretion (HOMA-B) for both Prediabetes and Normal Glucose tolerance (NGT) individuals (12-year Curve). A hyperbolics relationship was found to exist between HOMA-S and HOMA-B in both NGT and Prediabetes individuals. Though the Curve for prediabetes was shifted more toward the left and origin but the difference was less remarkable compared to 18 years.

**Figure 1C:**
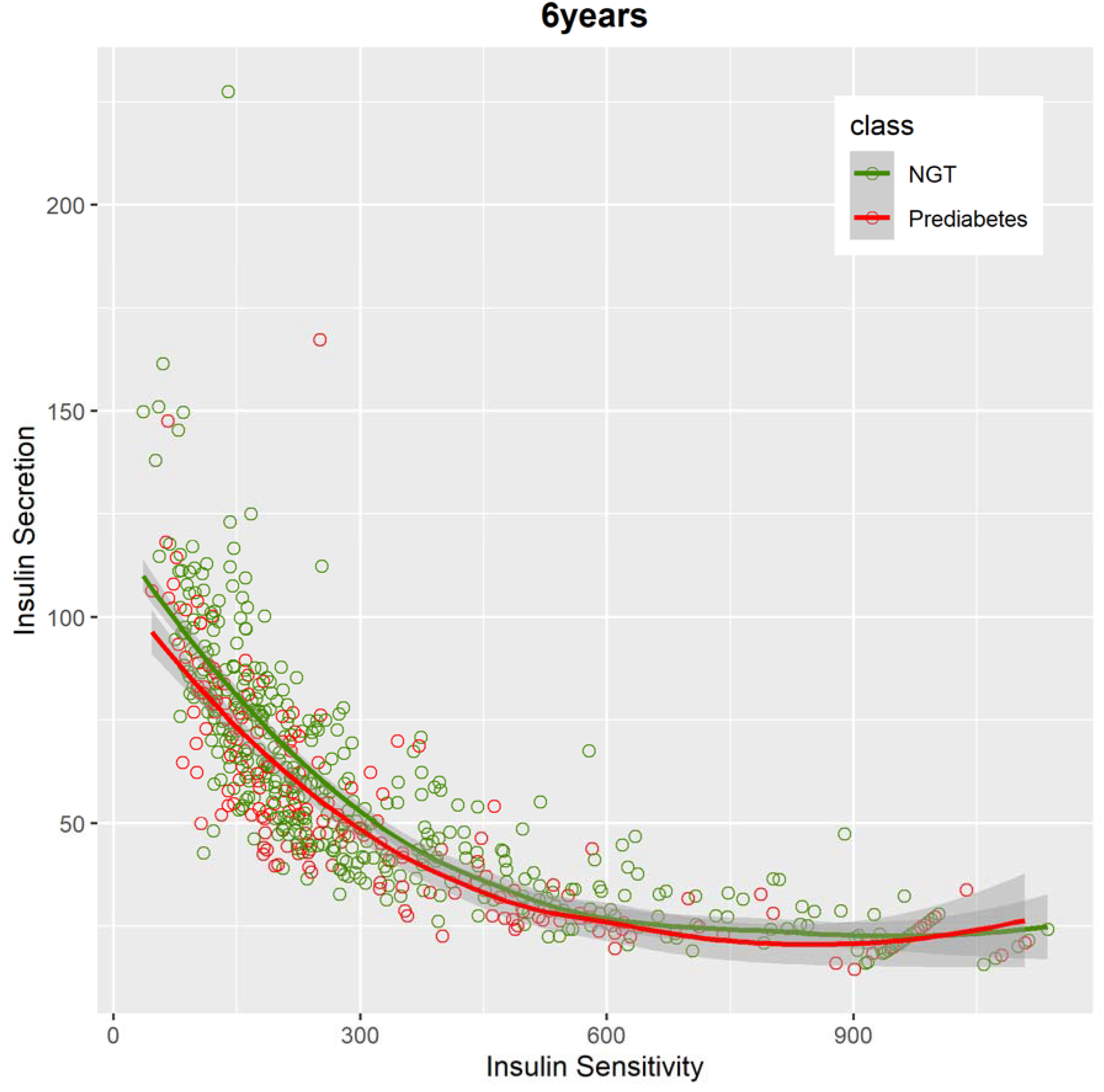
Relationship between Insulin Sensitivity (HOMA-S) and insulin secretion (HOMA-B) for both Prediabetes and Normal Glucose tolerance (NGT) individuals (6 Year Curve) A hyperbolics relationship was found to exist between HOMA-S and HOMA-B in both NGT and Prediabetes individuals. Though the Curve for prediabetes was shifted more toward the left and origin but the difference was less remarkable compared to 18 years.

### Trajectory of insulin sensitivity and β-cell function from 6 to 18 years

HOMA-S decreased and HOMA-β increased progressively in all participants. [Figure 2] Median HOMA-S fell by 39.2% between 6 to 12 years, and a total of 68% from 6 to 18 years. Median HOMA-β increased by 48.63 % from 6 and 12 years and 91.12% from 6 to 18 years. Disposition index reduced by 11% decrease between 6 to 12 years and 41.4 % decrease between 6 to 18 years.

**Figure 2:**
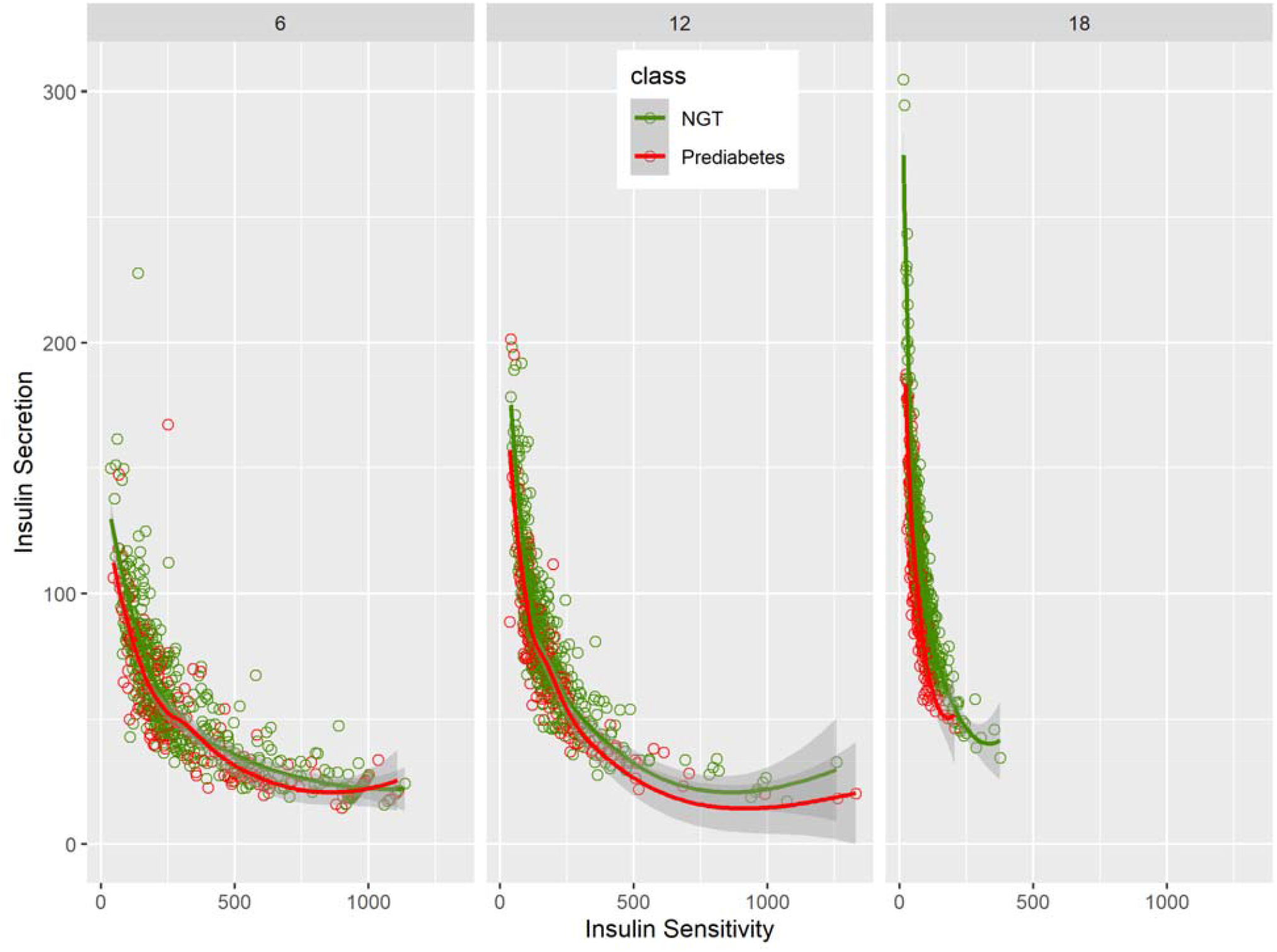
Evolution of Insulin Sensitivity and insulin secretion in NGT and Prediabetes from Childhood to Young adulthood. Over the years, Insulin sensitivity decreases in both NGT and Prediabetes. Decrease was more in prediabetes compared to NGT. Insulin secretion increases in both NGT and Prediabetes, increase was more in NGT compared to prediabetes.

The median decrease in HOMA-S between 6 to 18 year was 66.74% in NGT and 70.58 % in Glucose intolerant. The increase in HOMA-β between 6 to 18 years was 88.62 % in NGT and 91.48 % in glucose intolerant. The decrease in disposition index between 6 to 18 years was 39.5 % in NGT and 46.8% in glucose intolerant participants.

The life-course trajectories of HOMA-S in NGT and glucose intolerant overlapped considerably. On the other hand, the trajectory of HOMA-B in the glucose intolerant individuals started lower at 6 years and followed a lower path compared to the NGT throughout, though confidence intervals did overlap. Visualization of concurrent HOMA-B and HOMA-S at 6,12 and 18 years in the NGT and glucose intolerant groups showed that the latter group started with lower insulin sensitivity and beta cell function at 6 years and, continued to follow a lower trajectory throughout. The smoothened mean trajectories of the disposition index confirm the observation. [Figure 3]

**Figure 3:**
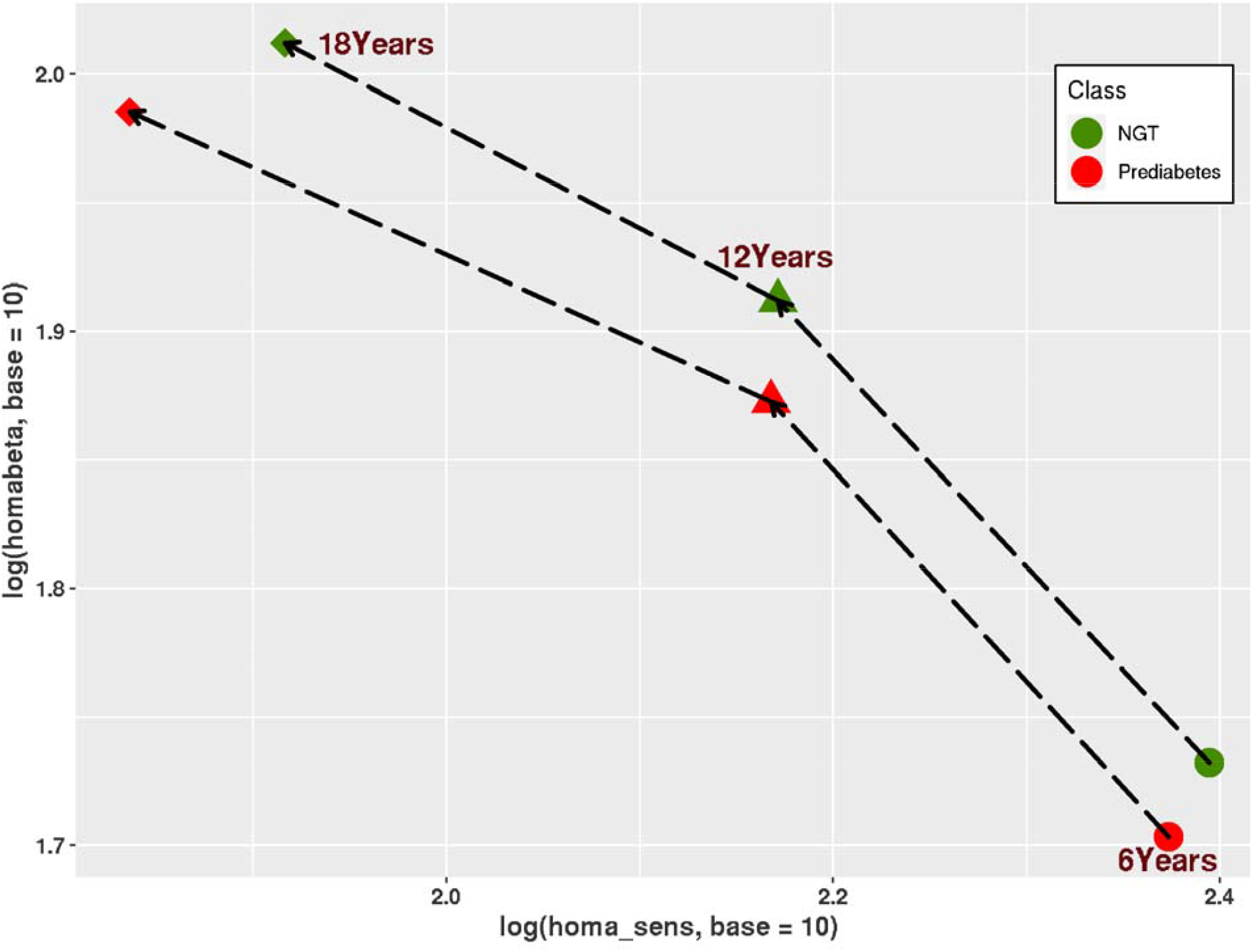
Median Trajectory of NGT and Prediabtes group. The curves represents median trajectory of disposition index for NGT and Prediabtes starting from 6 years to 18 years.

On linear mixed effect modelling, intercepts for both HOMA-S (471 in Glucose intolerant vs 492 in NGT, p=0.49) and HOMA-B (31 in Glucose intolerant vs 33 units in NGT, p=0.55) did not differ significantly. The intercept for the disposition index was significantly lower in the glucose intolerant group (17.5 × 10^3^ in Glucose intolerant vs 18.6 × 10^3^ in NGT, p=0.02), though the rate of change was not significantly different between the two groups [Supplementary information].

ROC curve analysis showed that the area under the curve was higher for the disposition index compared to both HOMA-S and HOMA-B at 6,12 and 18 years of age. The AUC for disposition index progressively increased with age and was 0.862 at 18 years of age. [Figure 4]

**Figure 4:**
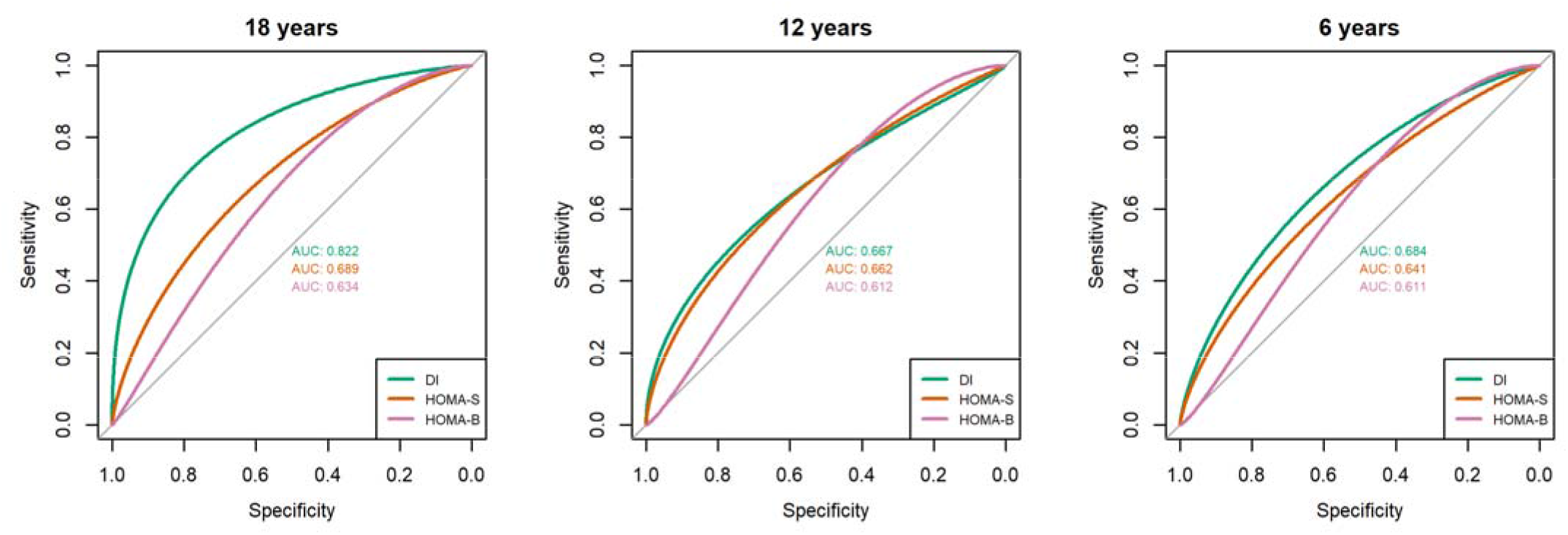
ROC curves for prediction of diabetes status (NGT vs Prediabetes) using HOMA-S, HOMA-B and Disposition Index at 6, 12 and 18 years timepoint.

## Discussion

We confirmed a hyperbolic relationship of insulin sensitivity and insulin secretion, both assessed using HOMA models, in a birth cohort of young Indians from childhood through young adulthood. Nearly one-third of these participants had glucose intolerance at age 18, despite a normal BMI. Bergman curves in those who were to be glucose intolerant at 18 years showed leftward shifts as early as six years. Longitudinal trajectories demonstrated that insulin sensitivity reduced and secretion increased with age in all participants, and taken in isolation, could not distinguish the NGT from the glucose intolerant. However, unlike those who retained NGT, the glucose intolerant did not manifest an increase in insulin secretion commensurate to the insulin insensitivity, which was captured by a lower trajectory of disposition index throughout early life.

The non-linear inverse relationship between insulin sensitivity and secretion implies an approximately constant product of these two parameters: a reduction in insulin sensitivity is accompanied by an increase in insulin secretion, thereby maintaining physiologic ability to dispose of glucose.[13] Though the original investigation used euglycemic hyperinsulinemic clamps and the intravenous glucose tolerance, the hyperbolic nature of the relationship has since been demonstrated across a variety of techniques of measurement, including using HOMA models [14]. In our cohort, while the hyperbolic curve was observed at all time points, serial measurements showed interesting shifts in the characteristics of these curves, showing an overall reduction in insulin sensitivity and increase in secretion. In addition, the curve shifted leftward-denoting inadequate physiological compensation to both reduced sensitivity and reduced secretion-as early as six years of age in those who would later have glucose intolerance.

Reduction in both insulin sensitivity as well as secretion is necessary for the hyperglycemia seen in diabetes to occur. Experimental insulin resistance (knockout of muscle insulin receptors) did not induce diabetes, but a knockout of both insulin receptors and IRS-1 (leading to beta-cell defect), resulted in diabetes, confirming the ‘2-hit’ paradigm. [15] Which one of these is affected first is contentious. Traditionally it has been felt that insulin sensitivity is compromised first; diabetes is precipitated when insulin secretion becomes deficient later in the course. [4] We have previously described the importance of early insulin insensitivity in another study. [16] Constructing trajectories from longitudinal data allowed a visual depiction of the evolution of the pathophysiology. While there were early signals, trajectories of the reduction of insulin sensitivity did not show significant differences between NGT and glucose intolerant. However, there were some (though statistically insignificant) reductions in insulin secretions in the NGT from childhood, and the difference was more pronounced when this was taken in the context of prevailing insulin sensitivity. Mechanisms of incommensurate insulin secretion are unclear, especially since they seem to occur before glucose levels are high to cause glucotoxicity related pancreatic cell damage. Early-life secretory defects could possibly owe to smaller organ mass in general, and beta cell mass in particular, in this population with documented inter-generational undernutrition. The association of a smaller birth length and head circumference in the prediabetic individuals in this cohort argue for inadequacy in attaining vital organ mass in the pathogenesis of the early onset diabetes in this population. [10] It would also be interesting to construct similar trajectories in the different types of diabetes;[17] we speculate that early secretory defects might be prominent in the severely insulin deficient diabetes (SIDD) which is most prevalent in Indians [18]

The disposition index was better than both HOMA-S and HOMA-B in predicting future glucose intolerance. This underlines that insulin secretion and sensitivity are not independent processes, and the pathogenesis follows a ‘capacity-load’ model: an incommensurately low capacity (insulin secretion) for the load (insulin insensitivity) differentiates individuals who develop prediabetes. Data from Pima Indians also showed that early inadequacies in both insulin action and corresponding secretion heralded a transition from NGT to IGT, and both these defects worsened in the further transition from IGT to diabetes. [19]

Childhood markers of future diabetes risk are helpful in offering a window for primordial preventions on a population level. A consortium of large studies (the i3c) showed that childhood BMI and glucose may predict adult diabetes. [20] However BMI may be difficult to use in our population given smaller birth sizes; all participants in this cohort who had glucose intolerance had normal BMI.[10] Our analyses argue for the potential utility of measuring the disposition index in childhood for such predictions, although admittedly this was not the aim of this study.

Knowing that the DI trajectories converged in early adolescence would be useful in avoiding this time point for measurement. The reason for this similarity is unclear and is possibly an effect of puberty.

This study has the strength of representing a well-characterized cohort with a high rate of follow-up, representing a population with a high prevalence of glucose intolerance despite low BMI. All measurements have been done using uniform, standardized methods at all time points. Weaknesses includes the lack of a frequently sampled GTT results, our reliance on HOMA models for both secretion and sensitivity and thus both rely on fasting insulin-ideally, the methods of measurement would have been independent of each other in plotting the Bergman plots. In addition, the outcome is glucose intolerance and not diabetes-it is possible that some individuals may not progress to diabetes and may even revert to NGT, like was seen in the Pima Indian study [19]. Further follow up of this cohort would shed more light on progression to ‘harder’ end-points. HOMA measurements are relatively easy and will be available in a large number of studies, compared to IVGTT/clamps. Our results should invite cohorts with serial data to validate our findings.

In summary, we found early evidence of glucose-insulin dysregulation in young, thin Indians who developed glucose intolerance in young adulthood. The trajectories of glucose insulin parameters facilitate an instinctually appealing visual understanding of the evolution of glucose intolerance. Childhood disposition index could differentiate these individuals from those who retained NGT from early childhood and could possibly be used to predict future diabetes risk.

## Data Availability

All data produced in the study are available on reasonable request to Prof Chittaranjan Yajnik

## Acknowledgements

The authors are grateful to all study participants and their family members for cooperation over many years. The authors thank the late Professor D.J.P. Barker, Dr. B. Coyaji, and Dr. V.N. Rao for their support in establishing the PMNS. The authors thank Dr. L. Garda, Director of KEM Hospital Research Centre. We also thank the staff of the Diabetes Unit for their help in conducting the study, particularly Drs. S. Hirve, N. Joshi, U. Deshmukh.; and H. Lubree, R. Ladkat, N. Memane, C. Joglekar, S. Bagate, A. Bhalerao, S. Chaugule, R. Dendge, T. Deokar, M. Gaikwad, N. Gurav, S. Jagtap, J. Kalokhe, S. Pandit, F. Rajgara, D. Raut, L. Ramdas, M. Raut, R. Saswade, and V. Solat. The authors thank Dr. S.S. Naik, Head of Biochemistry, KEM Hospital, for assay standardization.

## References

1. DeFronzo RA. Pathogenesis of type 2 diabetes mellitus. Med Clin North Am. 2004 Jul;88(4):787–835

2. Bergman RN, Ider YZ, Bowden CR, Cobelli C. Quantitative estimation of insulin sensitivity. Am J Physiol. 1979 ;236:E667–77.

3. Reaven GM, Hollenbeck CB, Chen YD. Relationship between glucose tolerance, insulin secretion, and insulin action in non-obese individuals with varying degrees of glucose tolerance. Diabetologia. 1989 Jan;32(1):52–5.

4. Bogardus C, Lillioja S, Howard BV, Reaven G, Mott D. Relationships between insulin secretion, insulin action, and fasting plasma glucose concentration in nondiabetic and noninsulin-dependent diabetic subjects. J Clin Invest. 1984 ;74:1238–46.

5. Eriksson J, Franssila-Kallunki A, Ekstrand A, Saloranta C, Widén E, Schalin C, Groop L. Early metabolic defects in persons at increased risk for non-insulin-dependent diabetes mellitus. N Engl J Med. 1989 10;321:337-43.

6. Pimenta W, Mitrakou A, Jensen T, Yki-Järvinen H, Daily G, Gerich J. Insulin secretion and insulin sensitivity in people with impaired glucose tolerance. Diabet Med. 1996 ;13(9 Suppl 6):S33–6.

7. India State-Level Disease Burden Initiative Diabetes Collaborators. The increasing burden of diabetes and variations among the states of India: the Global Burden of Disease Study 1990-2016. Lancet Glob Health. 2018 ;6:e1352–e1362.

8. Golden SH, Yajnik C, Phatak S, Hanson RL, Knowler WC. Racial/ethnic differences in the burden of type 2 diabetes over the life course: a focus on the USA and India. Diabetologia. 2019;62:1751–1760.

9. Yajnik, C.S., The insulin resistance epidemic in India: fetal origins, later lifestyle, or both? Nutr Rev, 2001. 59(1 Pt 1): p. 1–9.

10. Yajnik CS, Bandopadhyay S, Bhalerao A, Bhat DS, Phatak SB, Wagh RH, Yajnik PC, Pandit A, Bhave S, Coyaji K, Kumaran K, Osmond C, Fall CHD. Poor In Utero Growth, and Reduced β-Cell Compensation and High Fasting Glucose From Childhood, Are Harbingers of Glucose Intolerance in Young Indians. Diabetes Care. 2021 Dec;44:2747–2757..

11. Rao S, Yajnik CS, Kanade A, Fall CH, Margetts BM, Jackson AA, Shier R, Joshi S, Rege S, Lubree H, Desai B. Intake of micronutrient-rich foods in rural Indian mothers is associated with the size of their babies at birth: Pune Maternal Nutrition Study. J Nutr. 2001 ;131:1217–24.

12. American Diabetes Association. 2. Classification and Diagnosis of Diabetes: Standards of Medical Care in Diabetes-2021. Diabetes Care. 2021;44(Suppl 1):S15–S33.

13. Bergman RN, Phillips LS, Cobelli C. Physiologic evaluation of factors controlling glucose tolerance in man: measurement of insulin sensitivity and beta-cell glucose sensitivity from the response to intravenous glucose. J Clin Invest. 1981;68(:1456–67.

14. Ahrén B, Pacini G. Importance of quantifying insulin secretion in relation to insulin sensitivity to accurately assess beta cell function in clinical studies. Eur J Endocrinol. 2004 Feb;150(2):97–104. doi: 10.1530/eje.0.1500097. PMID: 14763905.

15. Kahn SE. The importance of beta-cell failure in the development and progression of type 2 diabetes. J Clin Endocrinol Metab. 2001 Sep;86(9):4047–58. doi: 10.1210/jcem.86.9.7713. PMID: 11549624.

16. Yajnik CS, Shelgikar KM, Naik SS, Sayyad MG, Raut KN, Bhat DS, Deshpande JA, Kale SD, Hockaday D. Impairment of glucose tolerance over 10 years in middle-aged normal glucose tolerant Indians. Diabetes Care. 2003 Jul;26(7):2212–3. doi: 10.2337/diacare.26.7.2212. PMID: 12832342.

17. Ahlqvist E, Storm P, Käräjämäki A, et al. Novel subgroups of adult-onset diabetes and their association with outcomes: a data-driven cluster analysis of six variables. Lancet Diabetes Endocrinol. 2018;6(5):361–369.

18. Prasad RB, Asplund O, Shukla SR, et al. Subgroups of patients with young-onset type 2 diabetes in India reveal insulin deficiency as a major driver. Diabetologia. 2022 Jan;65(1):65–78.

19. Weyer C, Bogardus C, Mott DM, Pratley RE. The natural history of insulin secretory dysfunction and insulin resistance in the pathogenesis of type 2 diabetes mellitus. J Clin Invest. 1999 ;104:787–94.

20. Hu T, Jacobs DR Jr, Sinaiko AR, et al. Childhood BMI and Fasting Glucose and Insulin Predict Adult Type 2 Diabetes: The International Childhood Cardiovascular Cohort (i3C) Consortium. Diabetes Care. 2020 ;43:2821–2829.

